# PP mediated the relationship between METS-IR and AIS in non-diabetic patients

**DOI:** 10.1101/2024.09.26.24314461

**Authors:** Zhaoyang Li, Ruirui Jia, Dong Chen, Hui Wang, Youyou Zhang

**Author notes:** **First author’s name:** Li Zhaoyang. **Corresponding authors**: Zhang Youyou.

## Abstract

**Background:** The correlation between metabolic score for insulin resistance (METS-IR) and high insulin normal glucose clamp test is stronger than other non-insulin-based insulin resistance scores. Insulin resistance was related to the occurrence and development of hypertension and stroke, however, the association between METS-IR and hypertension, as well as ischemic stroke is lack of evidence. This study aimed to determine the association between METS-IR and acute ischemic stroke in non-diabetic patients, and the mediating role of blood pressure parameters on it.

**Methods:** This hospital-based study included 557 non-diabetic subjects diagnosed with premature acute ischemic stroke. Logistic regression models and mediation analyses were conducted to assess the role of blood pressure parameters (systolic blood pressure, diastolic blood pressure, pulse pressure and mean arterial pressure) in the associations between METS-IR and risk of acute ischemic stroke.

**Results:** After adjusting for potential covariates, each SD increment of METS-IR was associated with 26% higher odds (OR = 1.26, 95% CI: 1.06-1.52) increment in ORs of acute ischemic stroke. The OR for acute ischemic stroke in patients with highest quartile of METS-IR was 1.73 hold (OR = 1.73, 95% CI: 1.12-2.70) higher than that in those with the lowest quartile of METS-IR. Furthermore, mediation analyses indicated that pulse pressure partially mediate the detrimental effect of insulin resistance (assessed by the METS-IR) on acute ischemic stroke (mediation percentage: 11.70%).

**Conclusions:** Our study suggests a relationship between METS-IR and acute ischemic stroke, and the association of METS-IR with acute ischemic stroke was partially mediated by pulse pressure.

## Introduction

According to the report from Global Burden of Diseases 2019 Stroke Collaborators, the absolute number of incident strokes increased by 70% and prevalent strokes increased by 85% from 1990 to 2019, respectively^1^. Meanwhile, stroke remained the second-leading cause of death and the third-leading cause of death and disability combined in 2019^1^. Furthermore, newest report from the World Stroke Organization emphasizes that global stroke mortality and disability-adjusted life years is projected to increase by 46.83% and 30.74% from 2020 to 2050, respectively^2^. Therefore, taking proactive preventive measures against stroke risk factors, especially controllable risk factors, is of great significance for reducing the global burden of stroke.

Insulin resistance (IR), refers to a complex condition of the body, in which the body experiences impaired insulin inhibition of liver glucose output and reduced insulin sensitivity in muscle and adipocyte tissues^3^ . Previous studies have explored the association between IR and stroke, but the results have been inconsistent. For example, an early systematic review showed that IR is a potential risk factor for stroke^4^, while a recent meta-analysis based on seven previous studies aimed to evaluate the association between homeostatic model assessment of insulin resistance (HOMA-IR) and stroke risk but did not find a significant association^5^. It is worth noting that this may be due to the relatively low sensitivity of the widely used HOMA-IR. For example, a recent study exploring the association between IR and cerebral small vessel disease (SVD) found that the association between Gutt insulin sensitivity index and SVD is more significant than the association between HOMA-IR and SVD^6^. Compared with the Gutt insulin sensitivity index that comprehensively consider peripheral insulin, liver insulin sensitivity, and the effects of body weight on peripheral tissue glucose uptake rate, HOMA-IR only reflects liver insulin sensitivity. In addition, in clinical practice, not all patients are subjected to insulin index testing upon admission. In summary, there is an urgent need to adopt more sensitive and cost-effective methods to obtain indicators reflecting IR.

Metabolic score for insulin resistance (METS-IR), a score defined as ln((2*fasting glucose)+fasting triglycerides)*body mass index)/(ln(high-density lipoprotein), has been found to have a better diagnostic performance to detect impaired insulin sensitivity compared to euglycemic-hyperinsulinemic clamp obtained from the intravenous glucose tolerance test^7^. And METS-IR was found to be significantly correlated with intravisceral, intrahepatic and intrapancreatic fat and fasting insulin levels^7^. Since its proposal in 2018, an increasing number of studies have explored the association between METS-IR and cardiovascular disease and found that METS-IR has stronger sensitivity and predictive value. For example, a prospective study of Korean adults without diabetes^8^ found that METS-IR was significantly positively associated with new ischemic heart disease. Similarly, a prospective study^9^ conducted among US adults showed that METS-IR is more significantly associated with all-cause mortality and cardiovascular mortality than HOMA-IR. However, currently, there is limited research exploring the association between METS-IR and acute ischemic stroke.

Previous studies indicated that the insulin sensitivity was associated with blood pressure levels or hypertension risk. For example, a meta-analysis based on 11 previous studies showed a significant positive association between fasting insulin levels or HOMA-IR and the risk of later hypertension^10^. Meanwhile, a large number of studies have confirmed that high BP is a dominant risk factor for stroke^11^). And several previous randomized controlled trials in diabetes patients showed that adequate control of blood pressure can effectively reduce the risk of CVD^12^, especially stroke^13, 14^. Overall, we speculate that that elevated BP may mediate the relationship between IR and stroke. A recent study of non-diabetic patients based on community residents showed that blood pressure partially mediates the association between IR and cerebral small vessel disease^6^, suggesting that in non-diabetes patients, blood pressure has a potential mediating role between IR and stroke.

In this study, we aimed to explore the association between METS-IR, blood pressure components (including systolic BP [SBP], diastolic BP (DBP), pulse pressure [PP] and mean arterial pressure [MAP] ) and the prevalence of acute ischemic stroke in non-diabetic patients. Meawhile, the potential mediating role of blood pressure components on the association between METS-IR and acute ischemic stroke were also detected.

## Methods

### Study participants

This was a hospital-based retrospective study, which was conducted between January 2023 and December 2023. Patients with stroke-like symptoms assessed by the brain computed tomography (CT) and diffusion-weighted imaging (DWI) scan were consecutively recruited from the department of geriatric neurology, the second affiliated hospital of Xi’an Jiaotong University. After further excluding the patients with venous thrombolytic therapy, cardiogenic cerebral embolism, hemorrhagic stroke, a history of diabetes, a history of either acute or chronic inflammatory or infectious diseases, chronic liver diseases, chronic kidney diseases, and neoplastic diseases. Also, we excluded individuals missing information on fasting blood glucose (FBG), lipid parameters and body mass index (BMI). Finally, a total of 557 patients were included in present analyses.

### Assessment of blood pressure and METS-IR

Blood pressures were measured using the electronic blood pressure monitor (Omron HEM7136, Japan) by trained staff, Also, anthropometric measurements including body height and weight were collected during the examination. Venous blood samples were drawn from the participants after fasting for at least 8-h. FBG, lipid parameters including total cholesterol (TC), triglyceride (TG), low density lipoprotein cholesterol (LDL-C) and high-density lipoprotein cholesterol (HDL-c) were detected at the clinical laboratory using automatic biochemical analyzer (Beckman AU5800, USA) . Pulse pressure (PP) was calculated computed as systolic blood pressure(SBP)-diastolic blood pressure(DBP) and mean arterial pressure(MAP) was computed as (SBP + 2× DBP)/3. BMI was computed as body weight(kg)/ body height(m)^2. METS-IR index was computed as ln[(2×FBG(mg/dl)+ TG(mg/dl))×BMI] / ln[HDL(mg/dl)].

### Assessment of covariates

Information of age, gender, and education level were recorded. Smoking or drinking status was categorized into ever and never smoking or drinking groups. Patients were considered smokers if they smoked on average more than 10 cigarettes per day for at least 1 year, and patients were considered drinkers if their average daily intake of ethanol was ≥30 mL for more than 1 years.

### Brain CT and DWI acquisition and analysis

All patients underwent brain CT (SIEMENS, China) and DWI (GE 3.0T, United States) scan after admission. Brain CT examination ruling out cerebral hemorrhage. High signal intensity in DWI is defined as acute ischemic stroke.

### Statistical analysis

Basic characteristics for continuous variables in normal distribution and categorical variables were calculated as mean±standard deviation and number (percentage), respectively. Variables in non-normally distributions were summarized as median (P25−P75). All the above variables were compared using the Student’s t-test or Mann-Whitney U test for continuous variables, and Chi-square test for categorical variables. Multivariate logistic regression analyses were employed to estimate the assciation between MET-IR and acute ischemic stroke. Furthermore, we examined whether the blood pressure components might mediate the association between MET-IR and acute ischemic stroke. Statistical analyses were performed using the R software (Version 3.6.3; R Core Team). A two-sided *P*-value<0.05 was considered statistically significant.

## Results

### Basic characteristics of study participants

Overall, the average age was 61.28 years for the included 557 participants, of which 63.0 % were male. Compared with those without acute ischemic stroke, acute ischemic stroke patients were more likely to be older, male, smoker, with lower levels of TC and HDL, while having higher levels of SBP, PP, MAP and METS-IR (**Table1**). No significant differences were observed for BMI, drink, education level, FBG, TG, LDL, and DBP.

**Table1.** Basic characteristics of study participants Characteristic.

| Characteristic | All<br>(n = 557) | Non-ischemic stroke<br>(n = 217) | Ischemic stroke<br>( n=340) | *P* * |
| --- | --- | --- | --- | --- |
| Age | 61.276 (12.195) | 59.604 (11.809) | 62.344 (12.333) | 0.009 |
| Gender |  |  |  | 0.005 |
| male | 351.0 (63.0%) | 121.0 (55.8%) | 230.0 (67.6%) |  |
| female | 206.0 (37.0%) | 96.0 (44.2%) | 110.0 (32.4%) |  |
| BMI (kg/m<sup>2</sup>) | 24.034 (3.303) | 23.801 (3.291) | 24.182 (3.307) | 0.184 |
| Smoke |  |  |  | <0.001 |
| Never-smoke | 437.0 (78.5%) | 187.0 (86.2%) | 250.0 (73.5%) |  |
| Current-smoke | 120.0 (21.5%) | 30.0 (13.8%) | 90.0 (26.5%) |  |
| Drink |  |  |  | 0.314 |
| Never-smoke | 480.0 (86.2%) | 191.0 (88.0%) | 289.0 (85.0%) |  |
| Current-smoke | 77.0 (13.8%) | 26.0 (12.0%) | 51.0 (15.0%) |  |
| Education |  |  |  | 0.144 |
| Primary school and below | 107.0 (19.2%) | 37.0 (17.1%) | 70.0 (20.6%) |  |
| Junior high school | 164.0 (29.4%) | 65.0 (30.0%) | 99.0 (29.1%) |  |
| High school | 158.0 (28.4%) | 55.0 (25.3%) | 103.0 (30.3%) |  |
| University or above | 128.0 (23.0%) | 60.0 (27.6%) | 68.0 (20.0%) |  |
| FBG | 5.100 (1.068) | 5.047 (0.881) | 5.134 (1.171) | 0.32 |
| TC | 4.071 (1.039) | 4.252 (1.042) | 3.955 (1.022) | 0.001 |
| TG | 1.495 (1.098) | 1.591 (1.486) | 1.434 (0.749) | 0.149 |
| HDL-c | 1.162 (0.286) | 1.233 (0.298) | 1.116 (0.269) | <0.001 |
| LDL-c | 2.573 (0.772) | 2.645 (0.775) | 2.526 (0.769) | 0.076 |
| METS | 36.688 (6.222) | 35.832 (6.367) | 37.234 (6.074) | 0.01 |
| SBP | 136.864 (22.224) | 133.470 (21.521) | 139.029<br>(22.425) | 0.004 |
| DBP | 84.596 (13.261) | 83.756 (12.438) | 85.132 (13.752) | 0.222 |
| PP | 17.423 (5.424) | 16.571 (4.929) | 17.966 (5.658) | 0.002 |
| MAP | 102.019 (14.935) | 100.327 (14.455) | 103.098<br>(15.156) | 0.031 |

\*: The continuous variables were compared by using t-test or Mann–Whitney U test, and the categorical variables were performed by chi-square test.
BMI: body mass index; DBP: diastolic blood pressure; FBG: fasting blood glucose; HDL-c:high density lipoprotein cholesterol; LDL-c: low density lipoprotein cholesterol; MAP: mean arterial pressure; METS: metabolic score for insulin resistance; PP: pulse pressure; SBP: systolic blood pressure; TC: total cholesterol; TG: triglyceride

### Associations between METS-IR and acute ischemic stroke

Each SD increment of METS-IR was associated with 26% higher odds (OR = 1.26, 95%*CI* : 1.06—1.52; Model 2; **Table 2**) increment in ORs of acute ischemic stroke after adjustment for covariates. In tertile analysis, highest tertile of METS-IR was associated with increased prevalence of acute ischemic stroke (OR = 1.73, 95% CI : 1.12-2.70, *P* _for trend_ = 0.014; **Table 2** ), when compared with those at the lowest tertile. In addition, we calculated the cutoff value of METS-IR and above cutoff (34.851) versus below cutoff of METS-IR was associated with a 59% higher risk of acute ischemic stroke (OR = 1.59; 95% CI : 1.11-2.29; **Table 2**).

**Table2.**
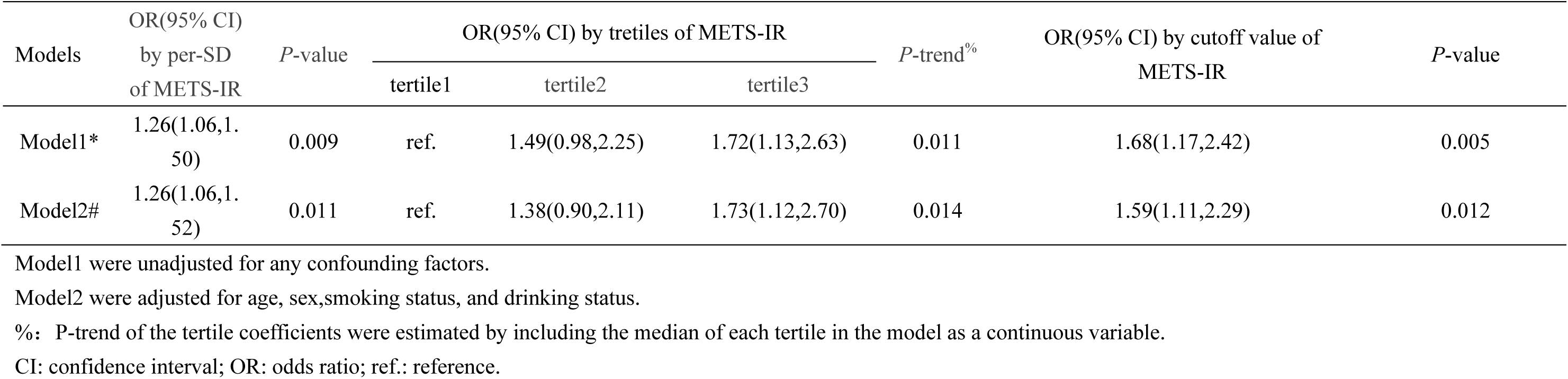
Associations between METS-IR and acute ischemic stroke.

| Models | OR(95% CI) | *P*-value | OR(95% CI) by tertiles of METS-IR |  |  | *P*-trend<sup>%</sup> | OR(95% CI) by cutoff value of | *P*-value |
| --- | --- | --- | --- | --- | --- | --- | --- | --- |
|  | by per-SD<br>of METS-IR |  | tertile1 | tertile2 | tertile3 |  | METS-IR |  |
| Model1* | 1.26(1.06,1.50) | 0.009 | ref. | 1.49(0.98,2.25) | 1.72(1.13,2.63) | 0.011 | 1.68(1.17,2.42) | 0.005 |
| Model2# | 1.26(1.06,1.52) | 0.011 | ref. | 1.38(0.90,2.11) | 1.73(1.12,2.70) | 0.014 | 1.59(1.11,2.29) | 0.012 |

Model1 were unadjusted for any confounding factors.
Model2 were adjusted for age, sex, smoking status, and drinking status.
%: *P*-trend of the tertile coefficients were estimated by including the median of each tertile in the model as a continuous variable.
CI: confidence interval; OR: odds ratio; ref.: reference.

### Associations between METS-IR and blood pressure parameters

After adjustment for confounders, higher METS-IR was associated with higher levels of all the blood pressure parameters (**Table3**). The analyses showed that each one-SD increase in METS-IR was associated with a 4.45, 2.45, 0.67 and 3.12 increase in SBP, DBP, PP and MAP, respectively.

**Table3.** Associations between METS-IR and blood pressure parameters.

|  | **β(95% CI) by per-SD of**<br>**METS-IR** | ***P*-value** |
| --- | --- | --- |
| SBP | 4.45(2.62,6.29) | <0.001 |
| DBP | 2.45(1.39,3.52) | <0.001 |
| PP | 0.67(0.24,1.09) | 0.00247 |
| MAP | 3.12(1.89,4.35) | <0.001 |

Models were adjusted for age, sex,smoking status, and drinking status.

### Associations between blood pressure parameters and acute ischemic stroke

With the exception of DBP, all other blood pressure parameters exhibit a significant positive correlation with acute ischemic stroke. Per one-SD increment in DBP, PP and MAP was associated with a 27%, 29% and 21% higher risk of acute ischemic stroke (all *P*<0.05; **Table4**).

**Table4.** Associations between blood pressure parameters and acute ischemic stroke.

| **Blood pressure parameters** | **$\beta$(95% CI) by per-SD of blood pressure parameters** | ***P*-value** |
| --- | --- | --- |
| SBP | 1.27(1.06,1.53) | 0.008 |
| DBP | 1.14(0.95,1.37) | 0.2 |
| PP | 1.29(1.07,1.57) | 0.008 |
| MAP | 1.21(1.01,1.46) | 0.034 |

Models were adjusted for age, sex, smoking status, and drinking status.

### Mediating role of SBP, PP and MAP in the associations between METS-IR and acute ischemic stroke

As shown in **Table5**, the significant mediated effects of PP on the associations between METS-IR and acute ischemic stroke were observed, with a 11.70% proportion mediated, whereas significantly mediated effects were not observed between SBP and MAP and acute ischemic stroke risk.

**Table5.** Assessment of mediating effects by blood pressure parameters on METS-IR and acute ischemic stroke.

| **Mediator** | **Total effect** | **Direct effects (95% CI)** | **Mediated effects (95% CI)** | **Mediated proportion (%)** | ***P*** |
| --- | --- | --- | --- | --- | --- |
| SBP | 0.007(0.001,0.01) | 0.005(-0.0002,0.01) | 0.001(0.0001,0.000) | 17.1 | 0.052 |
| PP | 0.007(0.002,0.01) | 0.006(0.001,0.01) | 0.01(0.0001,0.000) | 11.7 | 0.028 |
| MAP | 0.007(0.001,0.01) | 0.006(0.0001,0.01) | 0.001(-0.0003,0.000) | 13.3 | 0.134 |

Models were adjusted for age, sex,smoking status, drinking status.and corresponding intermediary variables(SBP, DBP, PP and MAP).

## Discussion

To the best of our knowledge, this is the first study to investigate the association between METS-IR and the prevalence of acute ischemic stroke among non-diabetic patients. Further mediation anaylsis suggested that PP would play a mediating role in the association between METS-IR and acute ischemic stroke.

Currently, research on the association between METS-IR and stroke is limited. A recent study involving patients with both hypertension and obstructive sleep apnea showed that each SD increase in METS-IR was associated with a 27% increased risk of incident stroke^15^. Another retrospective cohort study of hypertensive patients demonstrated a significant association between METS-IR and incident stroke, particularly ischemic stroke^16^. These results are similar to those found in our study, which indicates a significant positive association between METS-IR and ischemic stroke in non-diabetic patients. Considering the increasing number of individuals worldwide with impaired glucose tolerance and impaired fasting glucose, aside from those with diabetes^17^, the findings of this study highlight the importance for individuals without diabetes to be aware of the increased risk of stroke associated with insulin resistance. Prospective analysis needs to be conducted in a larger sample size to further validate the findings of this study.

An increasing number of studies have revealed the association between METS-IR and blood pressure. For example, a cross-sectional study involving 3,274 Chinese adults without hypertension showed a significant positive correlation between METS-IR and blood pressure values^18^. Another cross-sectional study based on 142,005 Chinese adults found a significant positive correlation between METS-IR and hypertension among those with normal weight^19^. The latest cross-sectional study involving 7,721 American adults also revealed a significant positive association between METS-IR and the risk of hypertension. Additionally, a study involving 15,453 Japanese adults found a significant positive correlation between METS-IR and the risk of prehypertension and hypertension among those with normal blood glucose levels^20^. These findings collectively suggest a strong association between METS-IR and blood pressure. In present study of a non-diabetic population, we found a significant positive correlation between METS-IR and blood pressure components, which once again demonstrates the close association between METS-IR and abnormal blood pressure.

Given that many studies have shown an independent link between abnormal blood pressure levels and ischemic stroke^11^, we further investigated the potential mediating role of blood pressure components in the relationship between METS-IR and ischemic stroke. Our results suggest a mediating role of PP in the association between METS-IR and ischemic stroke. PP is an index that calculated as the difference between SBP and DBP. A meta-analysis based on 11 previous studies showed that PP is an independent risk factor for stroke^21^. PP reflects the stiffness of the aorta and large arteries^22–24^. Meanwhile, similar to the significant positive correlation found between METS-IR and PP in our current research, two previous cross-sectional studies based on the US and Japanese adults also found a significant association between METS-IR and PP^25, 26^. Therefore, the potential mediating role of PP in the relationship between METS-IR and ischemic stroke identified in this study suggests that insulin resistance may increase the risk of ischemic stroke through the development of arteriosclerosis.

Several limitations need to be acknowledged when interpreting the results. Firstly, the diagnostic criteria for acute ischemic stroke in our study are based on clinical symptoms and signs, using DWI high signal as the imaging standard, which may have overlooked some acute ischemic stroke patients with negative DWI findings. Secondly, this is a retrospective, single-center, cross-sectional study, which failed to establish a causal relationship between the METS-TR, blood pressure components and ischemic stroke. Therefore, further validation of the causal relationships in a larger, multicenter cohort study is necessary, and the underlying mechanisms should be investigated through basic experimental studies in the future. Additionally, we were not able to adjust for some potential confounders such as the use of antihypertensive medication and physical activity, due to difficulty capturing accurate report of these confounders.

## Conclusions

Our study found that the association between insulin resistance measured by the METS-IR and acute ischemic stroke in non-diabetic patients. Furthermore, our results suggested that PP can partially mediate the association between the METS-IR with acute ischemic stroke. This study highlighted the value of METS-IR, a cost-effective indicator with high sensitivity, in predicting ischemic stroke in non diabetes patients, and preliminarily offered insights into the underlying mechanisms linking METS-IR and stroke by by revealing the mediation role of PP and METS-IR and ischemic stroke.

## Author Contributions

Zhang Youyou and Li Zhaoyang: Conceptualization, Formal analysis, and Writing-Original draft preparation; Zhang Youyou, Jia Ruirui and Chen Dong: Data Curation; Wang Hui and Li Zhaoyang : Writing-Reviewing and Supervision.

## Availability of data and materials

All data generated or analyzed during this study are included in this article. Further enquiries can be directed to the corresponding author.

## Declarations

### Ethics approval and consent to participate

All procedures performed in this study were in accordance with the ethical standards of the institution and with the 1964 Helsinki Declaration and its later amendments or comparable ethical standards. This study was approved by the Ethics Committee of the Second Affiliated Hospital of Xi’an Jiaotong University (Approval No. 2022200). The data are anonymous, and the requirement for informed consent was therefore waived.

## Consent for publication

Not applicable.

